# Distinctive modularity and resilience of structural covariance network in first-episode antipsychotic-naive psychoses

**DOI:** 10.1101/2021.10.08.21264777

**Authors:** Madison Lewis, Tales Santini, Nicholas Theis, Brendan Muldoon, Katherine Dash, Matcheri S. Keshavan, Konasale M. Prasad

## Abstract

**Objective:** Divergent findings in structural brain alterations in psychoses suggest that often-observed regions may exist within a network of susceptible regions. We built structural covariance networks (SCN) of volumes, cortical thickness, and surface area using the Human Connectome Project atlas-based parcellation of 358 regions on structural MRI data from 79 first-episode antipsychotic-naive psychosis patients (FEAP) and 68 controls.

**Methods:** Using graph theoretic methods, we obtained representative graph metrics of integration, segregation, resilience, centrality, importance, community structure, and hub distribution for all 3-morphometric features. We compared networks for resilience by simulated removal (“attacks”) of nodes and hubs, and simulated replacement of correlations among nodes in FEAP with that of controls using DeltaCon metric.

**Results:** Volume and thickness SCNs of FEAP showed differences in many graph metrics in opposite directions compared to controls but not surface area SCN. Network resilience did not show differences in the replacement simulation and hub “attacks, but betweenness centrality-based node “attacks” showed FEAP network disintegrating with relatively fewer node removals with preserved global efficiency compared to controls. In FEAP, communities consisted of regions from different lobes and hubs were more distributed than in controls.

**Conclusions:** Our findings suggest decreased heterogeneity and less differentiated community structure of FEAP network that may not be attributed to illness chronicity and medications. Contribution of cortical thickness but not surface area suggests that disease/developmental processes may preferentially affect cortical thickness. Near-similar resilience of FEAP and control networks may shield controls from developing illness but may prevent fuller long-term recovery in FEAP.

## Introduction

Schizophrenia is associated with widespread alterations in brain volume, cortical thickness, and surface area. Although morphometric differences are reported on nearly 50 different regions, the findings are inconsistent^1,2^ that are often attributed to heterogeneity of illness, stage of the illness and medication status. However, it may also be due to altered morphometry of different brain regions within a network showing different degrees of covariations among other region such that cross-sectional examination allows identifying a few regions to be showing group differences at a given point in time. Structural covariance networks (SCN) of brain regions can provide clues to such a possibility. SCNs are quantitative mathematical representations of covariation of regional volume, cortical thickness, and surface area. Between-subject variability of regional volumes being greater than between-subject differences in whole brain volume and between-subject differences in one region covarying with between-subject differences in other regions has been repeatedly observed^3,4^. Thus, SCNs can potentially highlight between-subject differences in regional morphometrics covarying with between-subject differences in other brain regions that cannot be tested by between-group comparisons of regions-of-interest (ROI). These covariance patterns have revealed structural ‘connectivity’ differences in schizophrenia compared to controls^5^ globally and regionally in relation to specific cognitive networks^5,6^. Although the SCNs do not reflect underlying white matter tracts, white matter tracts tend to correspond to the SCN edges^7^.

Existing studies have used atlases, e.g., the Desikan-Killiany-Tourville (DKT)^8^ and the Automated Anatomical Labeling (AAL)^9,10^ atlases that are based on anatomical landmarks or cytoarchitectonic patterns that may not reflect structural or functional connectivity. We used the Human Connectome Project (HCP) Atlas which parcellates the cortex into 358 functionally connected regions allowing for better functional/connectional interpretations of the results that can provide meaningful data on covariance structure of the regions. For example, Mitelman et al^11^ did not find covariance differences in whole thalamic volume with cortical Brodmann areas between schizophrenia and controls but when volumes of thalamic nuclei were included, patients showed correlation of pulvinar with frontal cortical BAs and in controls with centromedian nucleus^12^.

To our knowledge, this is the first study using the HCP atlas to examine SCN in FEAP using a graph theoretic approach. We calculated graph metrics of the SCNs for segregation (clustering coefficient and modularity), integration (characteristic path length and eccentricity), centrality (betweenness centrality), resilience (assortativity), and importance (degree). Besides, we identified network hubs (highly connected nodes) that may be important to the network structure and function. Since schizophrenia SCN has been reported to be less resilient compared to healthy counterparts^9^, we examined the FEAP SCN’s resilience compared to that of healthy controls (HC) using the SCN built on HCP-parcellated regions.

We examined 79 FEAP to minimize the impact of medication use and illness chronicity compared to 68 group-matched HC. Among the 13 studies that investigated the SCN of schizophrenia using graph theory^5,8,9,13-19^, 3 studies investigated the FEAP. One study reported higher degree nodes in patients and no difference between first-episode and chronic patients^8^. Another study reported no differences in pathlength and global efficiency over 6 weeks, but clustering coefficient, local efficiency and modularity being different at baseline but not at 6 weeks^20^. Wannan (2019)^16^ reported that first-episode schizophrenia SCN, but not chronic or treatment-resistant schizophrenia, showed significant covariance difference with HC in the subnetwork comprising of temporal and frontal regions.

We hypothesized that the FEAP SCN will show higher degree nodes, more hubs, and be less resilient compared to HC network based on previous studies that reported higher degree nodes^8,9^, higher number of hubs^10^ that were quantitatively different from that of controls^13^. Prior data on reduced network resilience in chronic schizophrenia patients^9^ has not been tested in FEAP. This is important because previous studies have shown increasing structural covariance strength as the disease progresses. Since the disease-related pathology starts well before the clinical manifestations, identifiable in the neonatal infants of schizophrenia patients^21^, we predicted that the resilience of the SCN of FEAP patients will be less than that of controls. Previous studies have examined one of the three morphometric features that could give an incomplete picture of the impact of disease/developmental processes on the network. In this study, we examined volume, cortical thickness, and surface area of HCP atlas-based parcellation.

## Methods

### Subject Recruitment

We recruited FEAP patients of both sexes between the ages 12 and 50 years diagnosed with schizophrenia, schizoaffective disorder, delusional disorder, psychotic disorder not otherwise specified, mood disorders per DSM-IV from inpatient and outpatient facilities of the Western Psychiatric Institute and Clinic, Pittsburgh. We excluded individuals with mental retardation, substance dependence within the past 6 months and/or substance abuse in the last month (both according to DSM-IV), significant medical/neurological disorders and prior antipsychotic treatment^22^. Consensus diagnosis was made by senior clinicians after reviewing the Structured Clinical Interview for DSM IV (SCID-IV) and medical records data, and follow-up of patients for about six months. The study was approved by the University of Pittsburgh Institutional Review Board. After complete description of the study, informed consent was obtained from all subjects.

### Imaging Methods

Details of MRI scanning are published^22^. Briefly, T1-weighted 3-dimensional spoiled-gradient-recalled (3D-SPGR) MRI data were acquired on a 1.5T GE whole-body scanner (124 contiguous coronal slices perpendicular to the anterior commissure-posterior commissure line, 1.5-mm thickness, steady-state pulse sequence: TE=5 msec, TR=25 msec, matrix=256×192, FOV=24 cm and flip angle=40°).

Using FSL 6.0, the images were motion and bias distortion-corrected, skull-stripped and then visually inspected for optimum segmentation and quality. The initial parcellation was performed using the DKT atlas^23^ on FreeSurfer 6.0. The cortical regions were further parcellated using the HCP atlas^24^, which consist of 180 cortical regions per hemisphere created based on a large sample of resting state functional connectivity data. The HCP parcellation was integrated into the FreeSurfer parcellation using the FreeSurfer’s fsaverage space^25^, and then mapped onto each individuals’ T1-weighted image space and had the volume, surface area, cortical thickness, and volume masks extracted for SCN construction. Good quality parcellation was successfully implemented on all scans.

### SCN analysis

Separate SCNs were built for the volume, surface area, and cortical thickness using partial correlation coefficients controlling for age, sex, and total brain volume using MATLAB. Additionally, a random graph was constructed from the group data by randomizing each edge 100 times. SCNs were intensity thresholded based on small worldness (σ) range to ensure that the networks were nonrandom by comparing the clustering coefficient and the pathlength of the SCN with the random SCN:

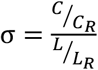

where *C* is the clustering coefficient and *L* is the pathlength^13^. Minimum threshold was the threshold at which both FEAP and HC graphs were nonrandom (σ>1.2). The threshold at which the graphs were not fully connected as determined by the reachability matrix was considered the maximum threshold.

At threshold intervals of 0.025 across the σ range, we calculated importance (degree), integration (characteristic path length and eccentricity), segregation (clustering coefficient and modularity), centrality (betweenness centrality), and resilience (assortativity) measures for the SCNs for each group for each morphometric measure using the Brain Connectivity Toolbox.^26^ Modularity, characteristic pathlength and assortativity were calculated as global measures across the σ range and others as nodal measures that were averaged to test for group differences using t-tests.

Hubs were defined as those nodes with degree, betweenness centrality, or eigenvector centrality being >2 standard deviations compared to network average^13^, and when found in ≥90% of the thresholded networks. All hubs were identified over the σ range at intervals of 0.005. Prior studies have defined hubs when they were found in 50% of the thresholded networks.

We, next, examined the modularity to investigate the community structure of the network at 21 different thresholds across the σ range as we did to identify the hubs.

#### Network Resilience

was examined using two methods: (a) simulated removal (“attacks”) of nodes without replacement in the SCNs of volume, cortical thickness, and surface area across the entire σ threshold range at 0.025 intervals. The giant connected component (GCC) and average efficiency of the network were measured following each simulated “attack”, and (b) replacement simulation using the DeltaCon metric^27^.

#### Removal simulations

First, we “attacked” the hubs arranged in the descending order of betweenness centrality and eigenvector centrality, separately, until all hubs were removed. But degree-based attacks were not implemented because all degree-based hubs were identified in the centrality-based simulations. Next, we sequentially “attacked” individual nodes in the descending order of degree, betweenness centrality and eigenvector centrality until no nodes remained in the network.

#### Replacement Simulation

The simulation was performed where the networks were the least similar across the σ range. The reference node was selected based on the lowest effect size in the case-control comparison of morphometry. The simulation adjusted the data for every node in the FEAP network such that their correlation with the reference node was equal to the corresponding edge in the HC network.

This “replacement” SCN was compared with the HC using the DeltaCon metric^27^, which is a single measure of network similarity that is more robust than other network comparisons. This single metric considers multiple graph properties, for example, the network impact of edge removals. This simulation was repeated five times and the results were averaged.

### Statistical Methods

We compared each morphometric measure for all regions separately for case-control differences using MANCOVA by including age, sex and total brain volume, followed by the Bonferroni-corrected between-subjects’ effects. T-tests were used to evaluate group differences in age, global graph measures across the thresholds, and for average nodal measures. χ^2^ test was used to compare sex distribution.

## Results

### Clinical and Demographic

Mean age and sex distribution of FEAP (23.99±7.21 years; 56 males) and controls (24.59±6.64 years; 39 males) did not differ (t=0.52, p=0.60; χ^2^=2.93, df=1, p=0.09). Mean illness duration from the time of first psychotic symptom to MRI was 2.57±3.35 years. FEAP included 47 schizophrenia, 13 major depressive disorder, 4 bipolar disorder, 3 delusional disorder and 12 psychotic disorder NOS subjects.

### Morphometric Comparisons

Seventy-three regions significantly different between groups in one or more of morphometric features. Two regions showed differences in all three morphometric features, namely left Area PGs (located at the superior surface of the angular gyrus), and right Area TE1 Middle. All group differences were of small effect sizes (Supplemental Table 1).

### Graph Measures

#### Graph metrics

For the global measures, assortativity was higher on average across the σ range for the volume SCN. For the volume SCN, all four nodal measures significantly differed. FEAP showed lower degree and higher clustering coefficient, betweenness centrality, and eccentricity. The cortical thickness SCN showed higher nodal values in all measures but betweenness centrality. For the surface area SCN, the only significant nodal measure was eccentricity and was dependent on threshold (Table 1, Fig 1).

**Table 1:** Average nodal and global graph measures across σ range for both groups for all three morphometric features.

| Average Graph Measure Across Threshold Range | Volume SCN |  | Cortical Thickness SCN |  | Surface Area SCN |  |
| --- | --- | --- | --- | --- | --- | --- |
|  | FB | HC | FB | HC | FB | HC |
| Global SCN metric |  |  |  |  |  |  |
| Characteristic Pathlength | $2.56 \pm 0.44$ | $2.42 \pm 0.37$ | $1.79 \pm 0.11$ | $1.84 \pm 0.11$ | $1.96 \pm 0.19$ | $1.97 \pm 0.18$ |
| Modularity | $0.44 \pm 0.10$ | $0.41 \pm 0.08$ | $0.16 \pm 0.04$ | $0.18 \pm 0.04$ | $0.28 \pm 0.07$ | $0.30 \pm 0.06$ |
| Assortativity | $0.31 \pm 0.04$ | $0.24 \pm 0.05$ | $0.10 \pm 0.03$ | $0.11 \pm 0.01$ | $0.23 \pm 0.06$ | $0.24 \pm 0.03$ |
| Nodal SCN Metric |  |  |  |  |  |  |
| Degree | $21.7 \pm 9.86$ | $24.8 \pm 10.76$ | $81.9 \pm 31.70$ | $65.4 \pm 27.75$ | $59.6 \pm 22.07$ | $58.7 \pm 20.31$ |
| Clustering Coefficient | $0.42 \pm 0.09$ | $0.39 \pm 0.06$ | $0.48 \pm 0.04$ | $0.41 \pm 0.03$ | $0.47 \pm 0.01$ | $0.46 \pm 0.01$ |
| Betweenness Centrality | $556.8 \pm 157.21$ | $507.4 \pm 132.95$ | $298.7 \pm 52.77$ | $319.6 \pm 58.67$ | $343.5 \pm 67.80$ | $345.4 \pm 63.63$ |
| Eccentricity | $3.80 \pm 0.86$ | $3.50 \pm 0.62$ | $3.16 \pm 0.40$ | $2.75 \pm 0.38$ | $3.12 \pm 0.45$ | $2.99 \pm 0.21$ |

**Fig 1:**
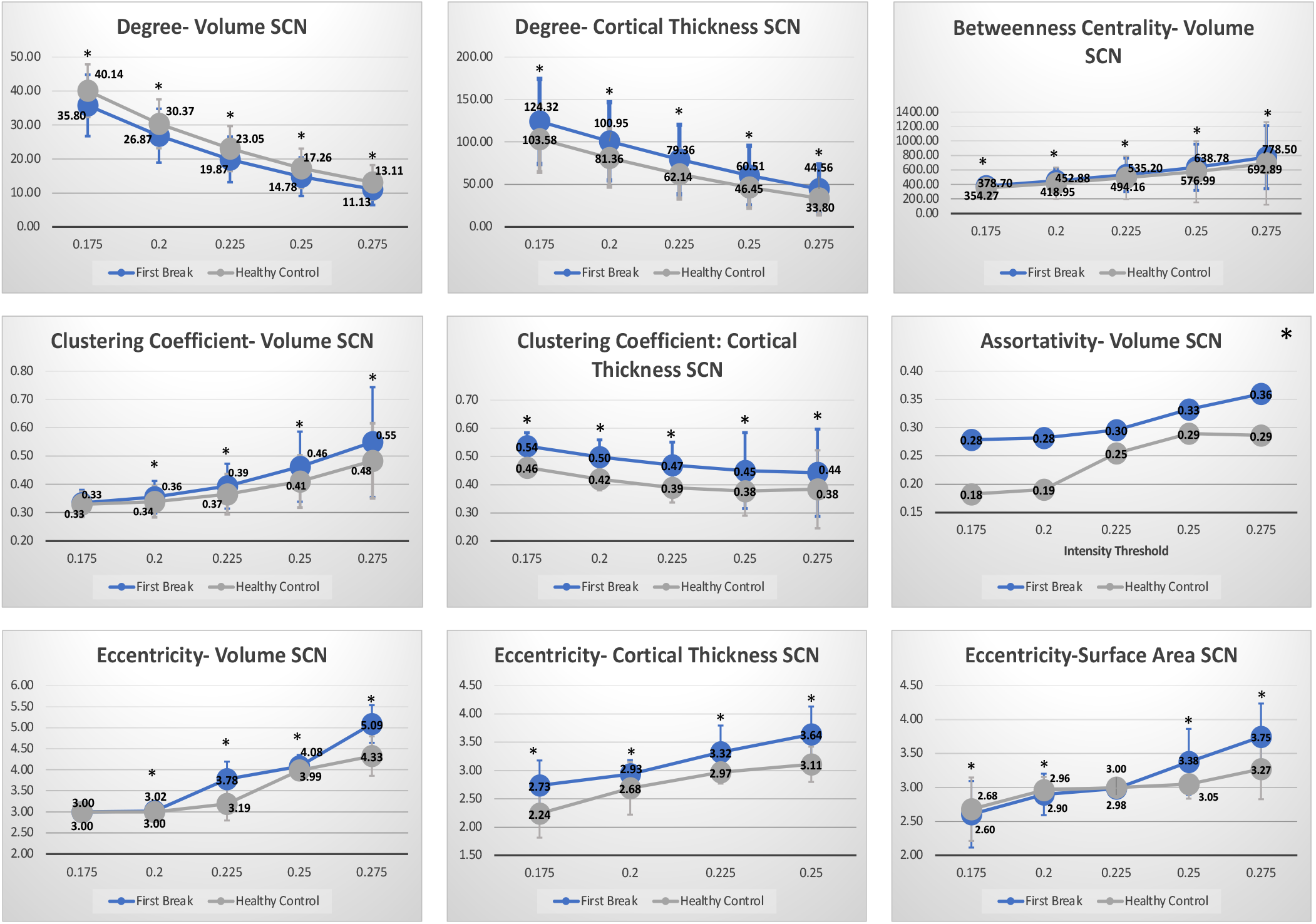
Global (top) and nodal (bottom) graph measures at intervals of 0.025 across the σ range for all morphometric features for FEAP (blue) and healthy control (gray). * represents significance between groups.

#### Hubs

Numerically higher number of hubs were found in the volume and surface area SCN of FEAP (Table 2), and were unique to each group in the volume, thickness, and surface area networks except for two in the surface area SCN. FEAP hubs were located in insular, frontal opercular, and auditory areas. The HC hubs were mostly located in visual, parietal, and insular areas (Fig 2).

**Table 2:**
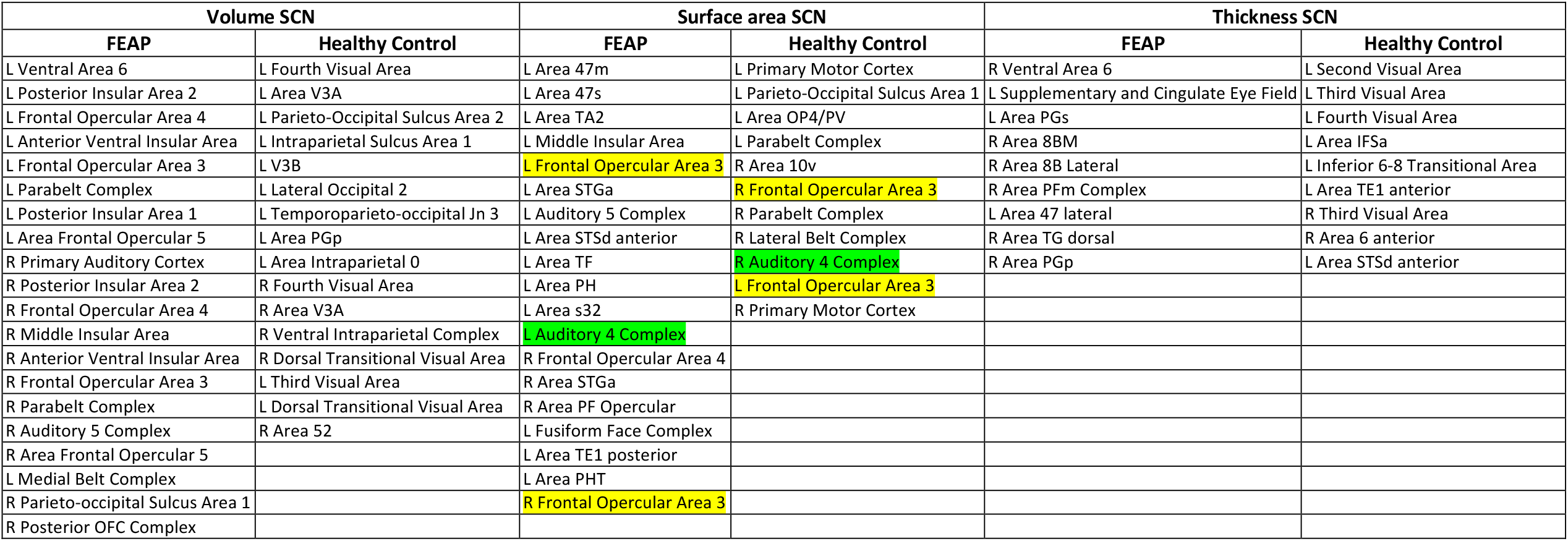
Regions identified as hubs in FEAP and controls in the volume, surface area and cortical thickness SCNs

| Volume SCN |  | Surface area SCN |  | Thickness SCN |  |
| --- | --- | --- | --- | --- | --- |
| FEAP | Healthy Control | FEAP | Healthy Control | FEAP | Healthy Control |
| L Ventral Area 6 | L Fourth Visual Area | L Area 47m | L Primary Motor Cortex | R Ventral Area 6 | L Second Visual Area |
| L Posterior Insular Area 2 | L Area V3A | L Area 47s | L Parieto-Occipital Sulcus Area 1 | L Supplementary and Cingulate Eye Field | L Third Visual Area |
| L Frontal Opercular Area 4 | L Parieto-Occipital Sulcus Area 2 | L Area TA2 | L Area OP4/PV | L Area PGs | L Fourth Visual Area |
| L Anterior Ventral Insular Area | L Intraparietal Sulcus Area 1 | L Middle Insular Area | L Parabelt Complex | R Area 8BM | L Area IFSa |
| L Frontal Opercular Area 3 | L V3B | L Frontal Opercular Area 3 | R Area 10v | R Area 8B Lateral | L Inferior 6-8 Transitional Area |
| L Parabelt Complex | L Lateral Occipital 2 | L Area STGa | R Frontal Opercular Area 3 | R Area PFm Complex | L Area TE1 anterior |
| L Posterior Insular Area 1 | L Temporoparieto-occipital Jn 3 | L Auditory 5 Complex | R Parabelt Complex | L Area 47 lateral | R Third Visual Area |
| L Area Frontal Opercular 5 | L Area PGp | L Area STSd anterior | R Lateral Belt Complex | R Area TG dorsal | R Area 6 anterior |
| R Primary Auditory Cortex | L Area Intraparietal 0 | L Area TF | R Auditory 4 Complex | R Area PGp | L Area STSd anterior |
| R Posterior Insular Area 2 | R Fourth Visual Area | L Area PH | L Frontal Opercular Area 3 |  |  |
| R Frontal Opercular Area 4 | R Area V3A | L Area s32 | R Primary Motor Cortex |  |  |
| R Middle Insular Area | R Ventral Intraparietal Complex | L Auditory 4 Complex |  |  |  |
| R Anterior Ventral Insular Area | R Dorsal Transitional Visual Area | R Frontal Opercular Area 4 |  |  |  |
| R Frontal Opercular Area 3 | L Third Visual Area | R Area STGa |  |  |  |
| R Parabelt Complex | L Dorsal Transitional Visual Area | R Area PF Opercular |  |  |  |
| R Auditory 5 Complex | R Area 52 | L Fusiform Face Complex |  |  |  |
| R Area Frontal Opercular 5 |  | L Area TE1 posterior |  |  |  |
| L Medial Belt Complex |  | L Area PHT |  |  |  |
| R Parieto-occipital Sulcus Area 1 |  | R Frontal Opercular Area 3 |  |  |  |
| R Posterior OFC Complex |  |  |  |  |  |

**Fig 2:**
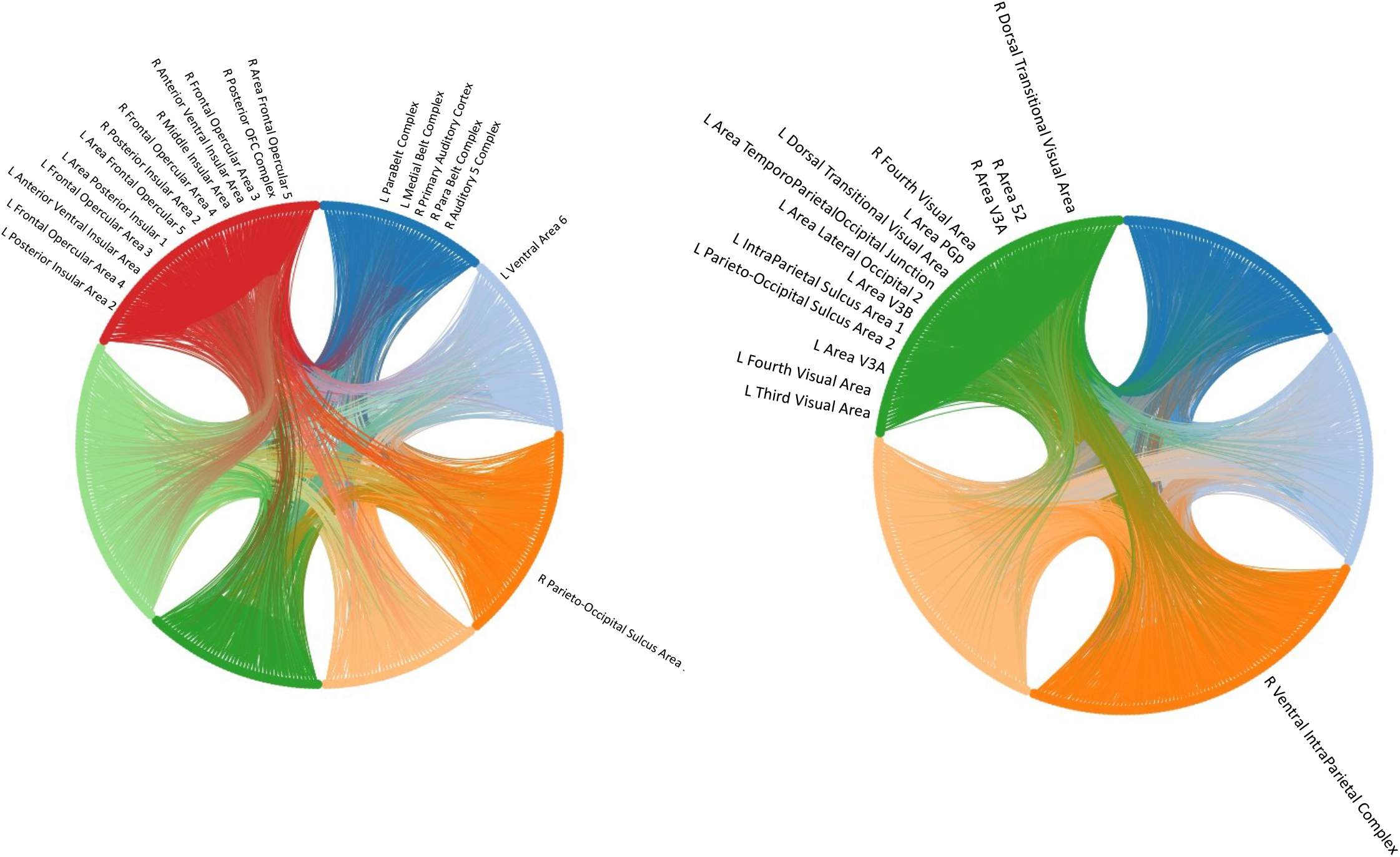
Circular graphs using NeuroMarvl (https://immersive.erc.monash.edu/neuromarvl/) showing modules at 0.225 threshold.

#### Modular structure

The number of modules increased with threshold across the σ range. There were seven modules in FEAP and five in control volume SCNs at 0.225 threshold (Fig 2). Surface area and thickness showed fewer modules (4/3 and 3/4, FEAP/HC). The modules comprised of minimally overlapping regions between the groups. FEAP modules consisted of auditory, limbic, and prefrontal nodes; HC modules comprised of visual areas (Supplemental Table 2, 3 and 4). Hubs were distributed across 4 communities in FEAP, but most hubs were in one community in controls.

### Network Resilience

Simulated “attacks” on hubs did not show group differences in network disintegration (Supplemental Figs 2&3). Betweenness centrality-based “attacks” disintegrated the GCC with 48.9% (175/358) of nodes removed for FEAP while 64.2% (230/358) nodes were removed for HC at a threshold of 0.275. At lower thresholds within the σ range, more nodes needed removal for network degradation in both groups. There were no differences when the threshold was 0.175. The efficiency did not show group-differences at any threshold tested (Fig 3; Supplemental Figs 4, 5 & 6). In the replacement simulation, the reference node for the volume SCN was the Right Insular Granular Complex. With replacement of the correlation values the similarity between groups increased minimally from 22.77% to 22.86% (Supplemental Fig 7).

**Fig 3:**
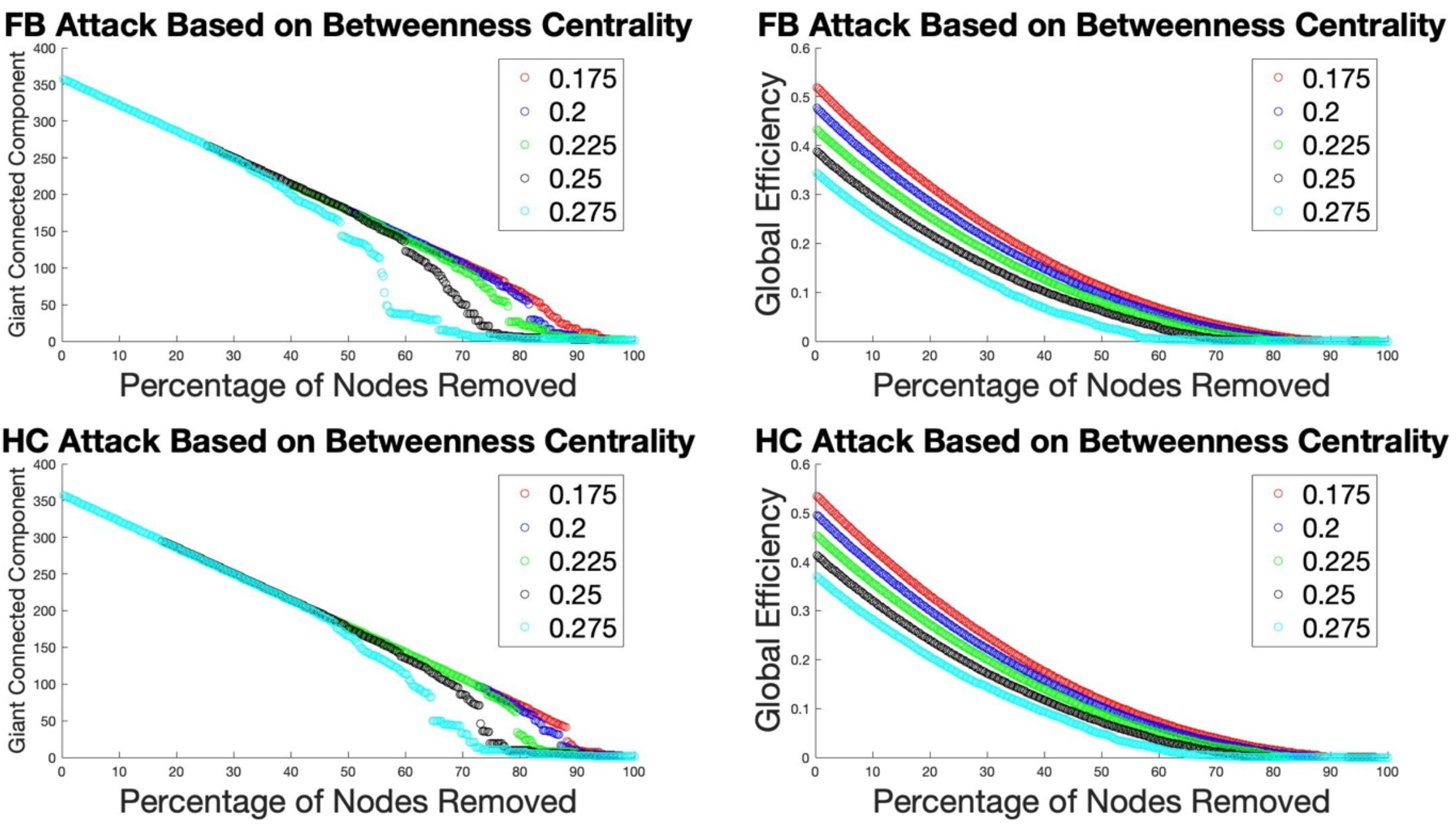
Network “attack” based on betweenness centrality using volume SCNs for both FEAP (top) and HC (bottom) across the σ range.

## Discussion

Our study is the first to use fine-grained, connectivity-based HCP parcellation to examine the SCNs of different morphometric features of FEAP and HC, and examine the network resilience using two different methods, to our knowledge. We find that the graph metrics were different in the SCNs of volume and cortical thickness but not of surface area. The FEAP SCN had longer pathlengths, greater tendency to form communities, higher likelihood of similar nodes connecting with each other, higher frequency of connecting with neighboring nodes rather than distant nodes, and the greater importance of nodes in the shortest paths compared to controls. There were more hubs and modules in patients compared to controls comprising of distinctly different regions in both groups. Modules comprised of regions from different brain lobes in FEAP compared to mostly from the same lobe in controls. Hubs were distributed across more communities in patients than in controls. Our simulations showed that the FEAP SCN may be more vulnerable to certain attacks, but not others with no difference in global efficiency suggesting that the FEAP and HC networks are nearly equally resilient. Thus, qualitatively, and quantitatively different SCN in FEAP may underlie some of the disease-related manifestations.

HCP-based atlas provides substantial details on connectivity-based parcellation providing a better context for studying the brain using a network approach than using traditional regional definitions that may not clearly depict underlying connectivity. To exemplify the degree of details that HCP parcellation provides, the area PGs as a hub that is common across morphometric features demonstrates functional connectivity to several areas within the lateral frontal, medial frontal, temporal, lateral parietal, and medial parietal lobes. Area PGs is structurally connected through the arcuate fasciculus/superior longitudinal fasciculus that courses anteriorly from PGs to inferior frontal sulcus and occipitotemporal junction parcellations. Local short association bundles connect with prefrontal and inferior parietal regions. Area PGs is involved in visuospatial attention, number processing, and response to biological motion, and redirects attention towards relevant stimuli^28^. Thus, examination of HCP-based examination can provide more fine-grained information on the connectivity pattern.

FEAP subjects showed higher global and nodal measures in volume SCN whereas cortical thickness SCN showed lower global, but higher nodal measures compared to controls. Low average degree in volume SCN suggests that the number of edges connected to each node in the FEAP SCN was lower than the number of edges on each node of the HC SCN. Since this is average nodal degree measurements, it is possible to have a few nodes with high degree with a majority of nodes having low degree. This is supported by the presence of more communities and hubs in FEAP compared to HC. Higher average clustering coefficient and betweenness centrality further supports the presence of several high degree nodes in the FEAP network than in HC SCN. Among other nodal measures, clustering coefficient, and betweenness centrality were higher in the volume SCN and showed a general trend in the same direction in the thickness SCN among FEAP compared to controls. Higher characteristic path length in FEAP suggests that the FEAP networks may have shifted away from random configuration compared to HC SCN^29^ and fewer interactions among neighboring nodes. Higher characteristic pathlength could introduce greater chances of network breakdown not allowing for more adaptive nodal interactions. Shorter pathlengths allowed greater chances of close physical interaction among proteins in protein-protein interaction network in schizophrenia.^30^ Thus, longer path lengths in FEAP SCN may prevent such interactions. Functional connectivity networks have also shown longer pathlength in schizophrenia^31^. Thus, investigation of multiple layers of networks simultaneously can provide greater insight into the network properties and functional implications.

The cortical thickness but not surface area SCN of FEAP was significantly different from HC SCN. Since volume is a product of surface area and thickness, our findings suggest that the disease/developmental processes may preferentially affect thickness, and that the contributions to volume-SCN may be through the cortical thickness rather than the surface area. Cortical thickness is related to laminar architecture and is highly conserved phylogenetically^32^. Existing data shows that the cortical thickness increases from early gestation period until early postnatal life^33^, increases by about 36% from birth to age 2 years when it nearly reaches its adult value, and then starts decreasing from 5 years of age in the sensorimotor areas first, followed by frontal lobe and then, the posterior parietal cortex until approximately 20 years of age.^34^ Cortical thickness changes are governed by cellular processes such as alterations in neuropil density, neuronal packing density, soma volumes, and myelination.^35^ Growth in surface area is closely tied to cortical curvature but driven by different processes. Cortical surface area continues to grow during childhood, achieving nearly 70% of its adult value by age 2^33^. Some region-specific cortical thinning has been observed in schizophrenia^36,37^ and in familial high-risk subjects who converted to psychosis^38,39^ that may be related to differential growth of inner and outer cortical layers,^40-43^ and is supported by cortical development during the evolution,^44^ and animal studies.^45^

Many outcomes of the graph measures are dependent on the threshold chosen. Small worldness-based threshold range provides a useful range to examine the network but does not provide one optimal threshold. Even the simulation-based tests for resilience were also dependent on the thresholding. This could be an issue when networks based on other modalities are tested together. We, recently, proposed a method that gives an optimal threshold and can be applied to networks built using imaging data across different modalities.^46^

Modular structure of FEAP SCN was distinctly different from that of HC SCN in that the FEAP modules comprised of regions from different lobes. The modular structure represents an optimized network organization^47^ for communication across spatially separated modules. Since the HCP atlas-derived regions are connectivity-based, FEAP modules may reflect a dysfunctional connectional pattern compared to control SCN that was more organized and tended to be representative of lobar organization. Such a modular organization may suggest underlying pathology related to disorganized thought processes and cognitive impairments.

Our finding of larger number of hubs in the FEAP SCN is not consistent with previous findings^9^, which may be due to more refined parcellation. Since the FEAP SCN is relatively sparsely connected, it may rely on more hubs to keep the network “functional.” In addition, larger number of hubs could indicate that the network may not be efficient and needs more edges or nodes to connect from one region to another. Many of these hub regions were morphometrically altered in prior studies^1^. Notably, none of the hubs in the FEAP SCN was in the heteromodal association area whereas many of HC SCN were in this region possibly suggesting that the FEAP SCN may be associated with large scale integration of multimodal information. Although some of the hubs for the FEAP are involved in higher level functioning tasks, the FEAP hubs lack the multimodal integration areas that were in the HC hubs.

Because the FEAP network is sparsely connected with more hubs, we predicted the network to be less resilient than HC SCN. Our observations partly supported our prediction. Both hub “attacks” and replacement simulation, evaluated by DeltaCon metric showed that the FEAP network was no less resilient than the HC SCN, which is inconsistent with a prior study that tested the network built on AAL-90 atlas^9^. Because of more refined parcellations, more nodes with higher betweenness centrality had to be removed in our study to make an impact on HCP SCN of FEAP. Betweenness centrality-based attack on nodes had the largest effect on network integrity suggests that nodes with high betweenness centrality may be vital for network integrity but not necessarily the hubs but the global efficiency did not change. It can be argued that the FEAP network may be more resilient to attack since the global efficiency was similar to HC SCN with the attacks even when network disintegrated. Replacement correlation showing minimal change in similarity of the FEAP network compared to the HC network suggests that the FEAP network may be fundamentally different from the HC network that may not be amenable to modification by just altering the correlational strengths between the nodes. Presence of resilience in FEAP and HC networks may serve different purposes in that it may provide resilience from developing psychosis in HC but resilience to remain symptomatic in FEAP. The latter may offer clues to why long-term recovery is about 14% even with antipsychotic treatment^48-51^.

There are several strengths in our study. We have examined FEAP that minimizes the impact of illness duration and medications providing a better estimate of neurobiological correlates. Examination of multiple morphometric measures simultaneously helps understand the SCN better. We have applied appropriate multiple test corrections where applicable, covaried for age, sex and total brain volume using partial correlation and examined the data for possible outliers. This study also uses two different types of simulations to examine network resilience more thoroughly. Limitations of our study includes modest sample size. Although imaging data were obtained on 1.5T scanner, the resolution was adequate to implement reliable morphometry. However, precise neurobiological impact is difficult to quantify in macroscale networks averaged across the groups. Emerging methods, such as structural similarity network (SSN)^39^ and individualized differential SCN^52^ analyses produce morphological networks for each subject that may allow one to meaningfully correlate with clinical measures.

## Data Availability

All data produced in the present work are contained in the manuscript.

## ACKNOWLEDGMENTS

This study was supported by the NIMH through R01 MH112584 and R01MH115026 (KMP); P50 MH045156, Behavioral Neuroscience and Schizophrenia; and NIH/NCRR/GCRC M01 RR00056 (Levine). We thank the faculty and staff of the Clinical Services Core of the Conte Center for the Neuroscience of Mental Disorders (P50 MH045156, Lewis) for their assistance in diagnostic and psychopathological assessments.

---

**Supplemental Table 1:** Morphometric differences in volume, cortical thickness and surface area between FEAP and controls of the regions that showed differences in at least two of the morphometric measure.

| Morphometric Comparisons- Multi-measure |  |  |  |  |  |  |  |  |  |  |  |  |  |  |  |  |  |  |
| --- | --- | --- | --- | --- | --- | --- | --- | --- | --- | --- | --- | --- | --- | --- | --- | --- | --- | --- |
| Region Name | Cortical Thickness |  |  |  |  |  | Volume |  |  |  |  |  | Surface Area |  |  |  |  |  |
| | FB Mean ± SD | HC Mean ± SD | Difference | ES | F | Sig. | FB Mean ± SD | HC Mean ± SD | Difference | ES | F | Sig. | FB Mean ± SD | HC Mean ± SD | Difference | ES | F | Sig. |
| Cortical Thickness, Volume, and Surface Area |  |  |  |  |  |  |  |  |  |  |  |  |  |  |  |  |  |  |
| L Area PGs | 2.62±0.21 | 2.68±0.17 | -2.28% | -0.33 | 4.57 | 0.03 | 2951.58±522.20 | 3215.93±535.80 | -8.22% | -0.50 | 14.59 | <.001 | 943.13±160.81 | 994.78±154.31 | -5.19% | -0.33 | 6.70 | 0.01 |
| R Area TE1 Mid | 2.61±0.32 | 2.71±0.26 | -3.63% | -0.34 | 4.59 | 0.03 | 1507.3±386.79 | 1669.68±345.16 | -9.72% | -0.44 | 12.94 | <.001 | 475.16±89.13 | 498.01±80.94 | -4.59% | -0.27 | 6.05 | 0.02 |
| Volume and Surface Area |  |  |  |  |  |  |  |  |  |  |  |  |  |  |  |  |  |  |
| L Sup 6-8 Trans A | | | | | | | 801.62±232.34 | 881.13±225.54 | -9.02% | -0.35 | 7.99 | 0.01 | 242.49±51.40 | 252.46±41.75 | -3.95% | -0.21 | 4.10 | 0.05 |
| R A TE1 post | | | | | | | 2910.16±594.02 | 3085.38±562.04 | -5.68% | -0.30 | 8.30 | 0.01 | 875.67±159.26 | 911.49±144.90 | -3.93% | -0.24 | 4.23 | 0.04 |
| R Sixth Visual A | | | | | | | 672.89±165.24 | 740.63±178.44 | -9.14% | -0.39 | 7.99 | 0.01 | 365.76±74.25 | 385.87±74.25 | -5.21% | -0.27 | 4.05 | 0.05 |
| Cortical Thickness and Volume |  |  |  |  |  |  |  |  |  |  |  |  |  |  |  |  |  |  |
| L A 3a | 1.87±0.11 | 1.91±0.10 | -1.77% | -0.32 | 6.45 | 0.01 | 1147.54±133.58 | 1189.24±125.36 | -3.51% | -0.32 | 6.05 | 0.02 | | | | | | |
| L A 47m | 2.64±0.32 | 2.73±0.35 | -3.40% | -0.28 | 4.11 | 0.04 | 409.22±102.11 | 427.59±86.22 | -4.30% | -0.19 | 4.29 | 0.04 | | | | | | |
| L Ventral A 6 | 3.07±0.29 | 3.17±0.30 | -2.96% | -0.32 | 4.64 | 0.03 | 991.78±216.26 | 1031.9±208.30 | -3.89% | -0.19 | 3.98 | 0.05 | | | | | | |
| L A Ant 32 prime | 2.74±0.19 | 2.84±0.19 | -3.44% | -0.52 | 13.75 | <.001 | 895.63±256.05 | 976.47±237.16 | -8.28% | -0.33 | 7.36 | 0.01 | | | | | | |
| L Inf 6-8 Trans A | 2.61±0.22 | 2.71±0.24 | -3.67% | -0.44 | 9.08 | 0.01 | 1078.41±252.64 | 1162.15±321.97 | -7.21% | -0.29 | 5.26 | 0.02 | | | | | | |
| L A TE1 post | 2.68±0.22 | 2.75±0.21 | -2.54% | -0.33 | 5.82 | 0.01 | 3427.46±618.30 | 3575.09±570.00 | -4.13% | -0.25 | 6.14 | 0.01 | | | | | | |
| R A OP2-3/VS | 2.72±0.23 | 2.79±0.25 | -2.34% | -0.27 | 4.17 | 0.04 | 428.49±58.04 | 443.29±77.15 | -3.33% | -0.22 | 4.60 | 0.03 | | | | | | |
| R Suppl & Cing Eye Field | 2.93±0.29 | 3.05±0.23 | -3.70% | -0.44 | 8.80 | 0.01 | 1533.77±274.76 | 1587.84±260.28 | -3.41% | -0.20 | 5.45 | 0.02 | | | | | | |

